# Diagnosing Others, Hiding Self: Shame and Non-Disclosure Among Autistic Psychiatrists - An Interpretive Phenomenological Analysis

**DOI:** 10.64898/2026.07.13.26357917

**Authors:** Mary Doherty, Nicholas Chown, Nicola Martin, Bernadette (Ret) Grosjean, Eddie Chaplin, Luna Dolezal, Sebastian C. K. Shaw

**Affiliations:** London South Bank University, UK; University College Dublin, Ireland; Harbor UCLA, USA; Department of Social and Political Sciences, Philosophy and Anthropology, University of Exeter; Department of Medical Education, Brighton and Sussex Medical School, University of Brighton and University of Sussex, Brighton, BN1 9PX

## Abstract

Autistic psychiatrists occupy a paradoxical position: trained to recognise and assess autism in others, yet navigating a professional culture in which their own autistic identity remains largely concealed. Despite growing visibility of autistic clinicians, the barriers autistic psychiatrists face to formal diagnosis and professional disclosure remain unexplored.

This study used interpretive phenomenological analysis to examine the experiences of seven autistic psychiatrists in relation to diagnosis and disclosure. Data were generated through in-depth interviews and Retzinger’s framework for identifying shame in discourse was applied as an analytical tool within the interpretive process.

Shame emerged as the overarching theme across the dataset, operating through four group experiential themes. Its origins lay in childhood experiences of difference and perceived defectiveness, transmitted through family, peers, and the broader social environment. In professional life, shame was sustained and amplified by colleagues’ misconceptions about autism, anticipated loss of credibility, and the deficit-based diagnostic criteria – which rendered self-recognition difficult and made formal diagnosis a perceived professional liability. Critically, shame did not only create barriers: it functioned as an override mechanism, rendering the known benefits of disclosure – to participants themselves, to colleagues, and to patients – insufficient to translate into action. This override function was not explained by fear of discrimination or rational career protection alone; it reflected shame’s operation as an internal prohibition, dissociated from its original social source and persisting even where stigma had been intellectually processed and rejected.

These findings reposition shame not as one barrier among many but as the organising force through which all barriers operate. Interventions aimed at increasing disclosure by raising awareness of its benefits misread the operative mechanism. Creating conditions in which autistic psychiatrists can make decisions about their identities freely requires naming and addressing shame – in research, in clinical training, and in the culture of psychiatry.

## Introduction

Autistic professionals are increasingly recognised across medicine,(1) education,(2) and the allied health professions,(3) among them individuals diagnosed both in childhood and in adult life.(4) This visibility challenges narratives that frame autism as incompatible with professional attainment, and raises important questions about what remains unseen: which autistic professionals exist, under what conditions they are recognised, and what sustains their invisibility.(5)

That visibility is, however, contested. Growing calls to restrict the autism diagnosis to those with complex needs or childhood presentation,(6–8) reflect a coordinated push along two fronts: formal proposals for a ‘profound autism’ subcategory defined by complexity and high support needs,(9) and emerging evidence that earlier- and later-diagnosed autism differ in genetic architecture, mobilised to question the validity of adult diagnosis.(10, 11) Yet the existence of autistic doctors – including those diagnosed in childhood – is itself evidence against the premise that these constitute distinct, non-overlapping groups. Frameworks that partition autism by complexity or developmental timing do not merely affect how autistic professionals are recognised: they perpetuate an incomplete and inaccurate understanding of autism, with consequences for autistic people across the full range of presentations.

Research into the experiences of autistic professionals has grown substantially in recent years. Autistic people are underrepresented in skilled employment relative to the general population, and autistic graduates are more likely to be unemployed than graduates with any other disability.(5, 12) Those who do enter professional roles in health and education face significant ongoing challenges, including discriminatory attitudes, inflexible institutional processes, and complex decisions about whether to disclose their autistic identity.(13) Disclosure carries considerable risk: autistic professionals report losing work opportunities, experiencing exclusion, and being subject to lowered professional expectations following disclosure.(14, 15) Many adopt masking strategies – performing neurotypicality at significant personal cost – to navigate workplaces that were not designed with them in mind.(16, 17) Late diagnosis is particularly common among autistic professionals, many of whom spent years developing sophisticated compensatory strategies before recognition, sometimes arriving at diagnosis only following burnout or a significant mental health crisis.(18)

Within medicine, psychiatry occupies a particular position with respect to autism. Psychiatrists are central to the diagnostic pathway for autistic people across many healthcare systems, and are uniquely placed to shape both clinical practice and broader cultural understanding of autism.(19) However, autistic identity remains largely invisible within the profession. Autism prevalence in the general population is estimated at approximately 1–2%, though this figure is likely a substantial underestimate; systematic analysis of primary care data has identified marked underdiagnosis, particularly in adults.(20, 21) Among doctors seeking mental health support, screening rates are considerably higher, with psychiatrists showing significantly elevated odds of positive screening compared with other specialties.(22)

Within Autistic Doctors International (ADI), psychiatry represents the second largest specialty group, suggesting autistic psychiatrists may be substantially over-represented relative to their proportion of the medical workforce.(4) Systematic prevalence data for the specialty nonetheless remain absent. The position of autistic psychiatrists is therefore paradoxical: trained to recognise and assess a neurotype they themselves may share, yet navigating a professional culture in which that identity remains largely undisclosed. This invisibility has implications not only for autistic psychiatrists themselves, but for the patients they treat and the clinical frameworks they apply.(23)

The decision not to disclose an autistic identity in professional settings is rarely straightforward. For autistic professionals across health and education, disclosure involves careful calculation of anticipated discrimination, access to support, professional reputation, and the costs of continued concealment.(5, 14) Stigma, both enacted and anticipated, functions as a powerful deterrent.(24) Within medicine, additional pressures pertain: concerns about fitness to practise, regulatory scrutiny, and cultures of professional stoicism that discourage disclosure of any personal difficulty.(4, 13) These structural and psychological pressures combine to ensure that autistic psychiatrists who do exist remain largely unknown to each other, to their institutions, and to the patients they serve.(23)

Without greater visibility of autistic professionals across the full diagnostic range, our understanding of what it means to be autistic, and of what barriers prevent that visibility from emerging, remains incomplete. Despite this, qualitative investigation of the lived experiences of autistic psychiatrists is limited,(25) and exploration of the barriers they face to diagnosis and to professional disclosure remains absent from the literature. This paper addresses that gap, examining two research questions: what factors shaped participants’ pathways to an autism diagnosis, and what influenced their decisions about professional disclosure?

## Methods

### Study design

This qualitative study uses interpretive phenomenological analysis (IPA), applied to the same dataset as a previously reported study of the experiences of autistic psychiatrists in relation to recognising autism.(25) IPA acknowledges and embraces the researcher’s insider perspective, prior knowledge and subjectivity in conducting the research and uses this to access experiences and data from participants which might not otherwise be obtained.(26) Retzinger’s (1995) framework for identifying shame in discourse was used as an analytical tool within this interpretive process.(27)

### Participants

Full details of participants, data collection and initial analysis are reported in the first paper from this dataset.(25) Briefly, participants were recruited via purposive sampling through the ‘ADI Psych’ WhatsApp group – a psychiatry-specific subgroup of ADI. Eligibility criteria were being a psychiatrist, not in training, and either self-identified as or was formally diagnosed as autistic. This paper reports seven participants. Eight were initially recruited; one was excluded after institutional review due to concerns about identification. Participant demographics are presented in Table 1.

**Table 1.** Participant demographics.

| Name | P1 | P2 | P3 | P4 | P5 | P6 | P7 |
| --- | --- | --- | --- | --- | --- | --- | --- |
| Age | 75-80 years | 40-45 years | 40-45 years | 35-40 years | 45-50 years | 40-45 years | 50-55 years |
| Gender | Male | Female | Female | Female | Female | Female | Male |
| Ethnicity | White / other | Indian | White British | White /other | White British | White / other | White / other |
| Employment | Retired | Full time | Full time | Full time | Part time | Part time | Full time |
| Experience in psychiatry | > 40 years | 10-15 years | 10-15 years | 10-15 years | 20-25 years | 15-20 years | 15-20 years |

### Data analysis

Data analysis followed the framework described by Smith and Nizza,(28) and was conducted in two distinct analytical phases. The first analytical phase involved, as reported previously, annotation of transcripts with exploratory notes, development of experiential statements, clustering into personal experiential themes (PETs) for each participant, and cross-case analysis resulting in group experiential themes (GETs).

The analysis presented in the first paper from this dataset focused on experiences of recognition.(25) The present analysis interrogates the same dataset in relation to two paired research questions: the reasons autistic psychiatrists sought or avoided formal diagnosis, and the reasons they chose to disclose or not disclose their autistic identity in professional contexts.

Qualitative analysis is inherently iterative; returning to data across multiple rounds is central to the method. Following completion of the initial analysis, and in the process of preparing the data for writing, the first author recognised shame as a possible overarching organising frame. This recognition was shaped by engagement with the shame and medicine literature and, specifically, by contact with co-author and shame theorist Luna Dolezal, who directed attention to Retzinger’s (1995) framework for identifying shame in discourse.(27)

In response, the first author returned to all included data sources – reviewing video recordings, audio, and transcripts – and conducted a further analytical phase using Retzinger’s indicators as a specific analytical tool within the interpretive process, consistent with the exploratory linguistic annotation described by Smith and Nizza.(28) This phase specifically attended to Retzinger’s three categories of shame indicator:

- *Verbal cues*: direct shame references and shame-related code words;
- *Verbal hiding behaviours*: mitigation, abstraction, denial, distraction, projection, and verbal withdrawal;
- *Paralinguistic and visual gestures*: over-soft speech, hesitation, self-interruption, laughed words, irregular rhythm, fragmented speech, and bodily manifestations of shame.

The identification of shame indicators across all three categories, occurring in combination, was taken as the strongest evidence for shame states, consistent with Retzinger’s guidance that convergent cues across categories provide more robust identification than single indicators.(27) This second pass through the analysis led to the creation of new GETs in addition to the alteration of existing ones. Independent verification of the analysis was provided by co-author NC.

### Reflexive participant collaboration

Reflexive participant collaboration was used.(29) Participants were offered the opportunity to review transcripts and confirm omission of potentially identifying data, and subsequently to read a draft paper and comment on the analysis and interpretation. Prior to formal dissemination, participants were also proactively notified of the emerging shame findings, in recognition of the potential emotional impact of the analysis and in keeping with the relational obligations of insider research. Five participants responded, with no changes to the analysis.

### Positionality

We are a mixed-neurotype research team. The first author is an autistic consultant anaesthetist and founder of ADI, the organisation through which participants were recruited. As detailed previously, this constitutes an ‘almost insider’ position: sharing autistic identity and medical professional identity with participants, but not the experience of psychiatry as a practising psychiatrist.(25) BG is an autistic psychiatrist and Psychiatry Lead for ADI. SCKS is multiply neurodivergent including autistic, and is ADI Research Lead.

The present analysis required additional reflexive consideration. Studying shame in a population to which one belongs – using data generated through relationships of trust – meant that the analytical process was not separate from the phenomenon under study. Shame, as Retzinger describes it, tends towards concealment; it hides from the person experiencing it as readily as from outside observers.(27) That it remained invisible across two prior analytical rounds is consistent with this, but it is also consistent with something else: that returning to this data, when the time came, carried a personal cost that existing frameworks for insider qualitative research do not adequately describe. The analysis presented here was not produced without difficulty. There were periods when the weight of what the data contained made continuation impossible – not because the findings were unclear, but because they were too close. The first author has written elsewhere about this experience.(30) It is documented here because it is methodologically relevant: the cost of insider research on shame is itself evidence of how shame operates.

The first author remained alert to the ways in which proximity to the phenomenon under study may shape interpretive choices – including the risk of over-identification with participants and the possibility that aspects of shared experience may be foregrounded or occluded. At the same time, the insider position was not incidental to the analysis: the recognition of shame as the overarching theme required a researcher for whom shame was not merely conceptual.

### Ethics

All procedures comply with the ethical standards of the relevant national and institutional committees on human experimentation and with the Helsinki Declaration of 1975, as revised in 2013. Ethical approval was granted by London South Bank University Research Ethics Committee; ETH2122-0128.

## Results

### Demographics

Seven participants were included in this analysis. Five were consultant psychiatrists and two were senior associate specialists. All were based in the UK. Six were currently employed and one was retired. Four were child and adolescent psychiatrists; the others were old age, liaison and general adult psychiatrists. One had a formal autism diagnosis received in adulthood. One had been diagnosed with pervasive developmental disorder not otherwise specified (PDD-NOS) in childhood and identified as autistic as an adult. The remaining five self-identified as autistic as adults. Participant numbers have been used and identifying details changed or omitted to preserve anonymity. See Table 1 for participant demographics.

### Results Summary

Four group experiential themes were identified, reflecting both the origins of shame and its operation across participants’ professional and personal lives. The second analytical phase produced two new group experiential themes: GET 1 (shame permeated everything) and GET 2 (origins of shame) – in addition to refining and re-interpreting two pre-existing ones: GET 3 (shame in professional life) and GET 4 (shame as override). See Figure 1.

Shame pervaded participants’ experiences as an overarching organising theme, present in both overt and bypassed forms across the dataset (GET 1). Its roots lay in childhood and early life – in messages absorbed from family, peers, and the social environment that encoded autism as fundamental deficiency long before professional training compounded them (GET 2). On entering and navigating psychiatric practice, shame shaped the professional environment: driving concealment, generating barriers to diagnosis and disclosure, and finding structural reinforcement in a specialty whose deficit-based frameworks participants had absorbed and applied (GET 3). Critically, shame did not only create barriers – it rendered the incentives for disclosure insufficient, overriding the capacity to act even when participants could clearly articulate what disclosure might offer them, their colleagues, and their patients (GET 4). These themes do not operate in isolation. The shame that originated in childhood was carried into professional life; the professional environment reactivated and amplified it; and its override function foreclosed the very actions that might have interrupted the cycle.

Within each GET, sub-themes are described as follows. GET 1 comprises two sub-themes: overt shame in discourse (1a) and bypassed shame (1b). GET 2 comprises four sub-themes: the self as defective (2a); family and early messages (2b); school and peers (2c); and generational shame (2d). GET 3 comprises four sub-themes: lack of knowledge, myths and misconceptions (3a); professional identity and credibility under threat (3b); diagnostic criteria and internalised ableism (3c); and navigating the environment – individual strategies (3d). GET 4 comprises three sub-themes: the benefits of visibility – present but insufficient (4a); the advocacy pull and its cost (4b); and reasonable accommodations as anticipatory shame (4c). The override mechanism – shame’s capacity to render known benefits of disclosure insufficient to translate into action – is the organising finding of GET 4.

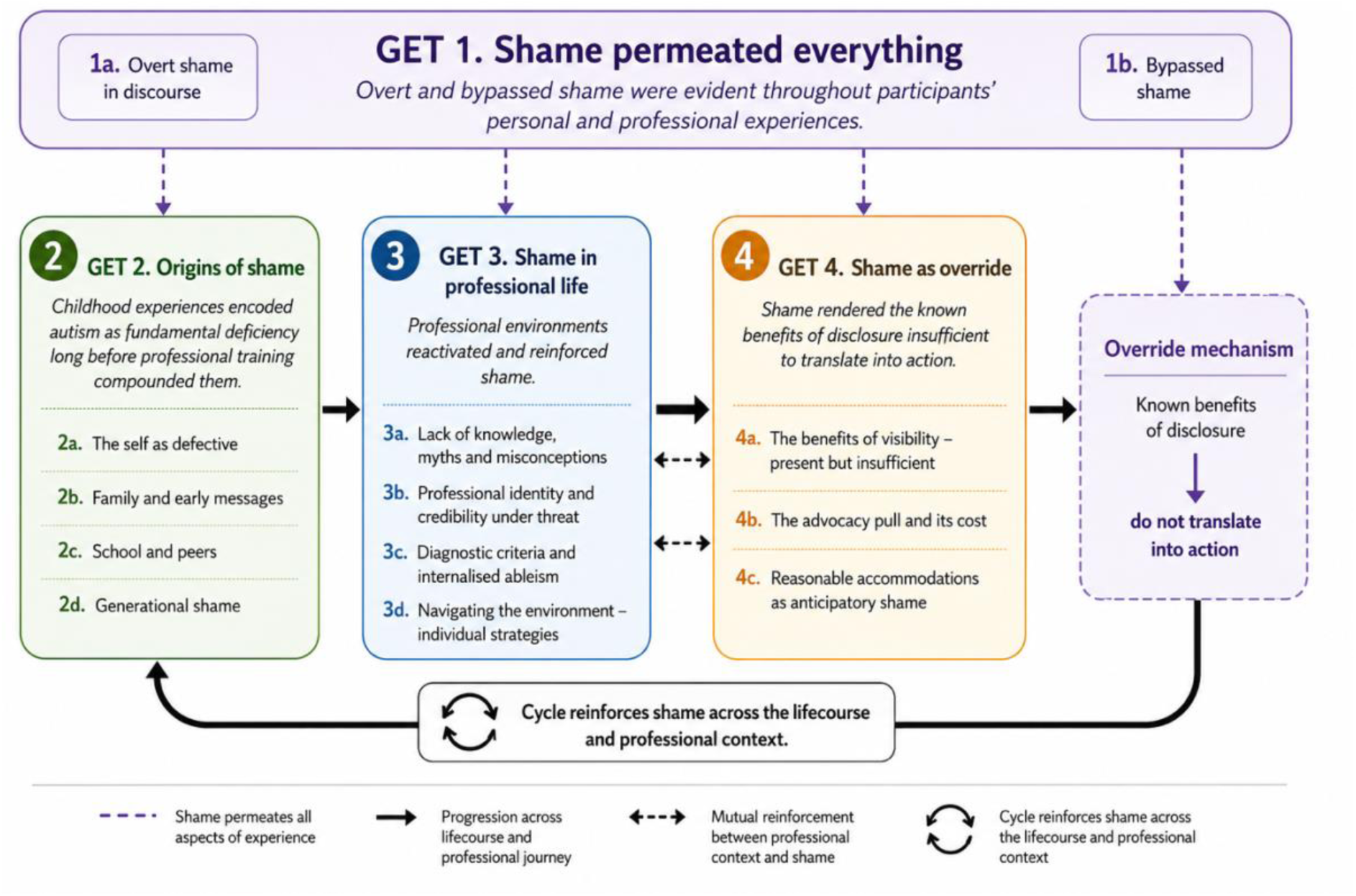

### GET 1: Shame permeated everything

Analysis of participants’ experiences revealed shame as an overarching theme operating across two distinct but related areas: the pathway to formal diagnosis, and decisions about whether to disclose an autistic identity professionally.

This dataset had previously been analysed in relation to identity and recognition,(25) and subsequently for this paper in relation to diagnosis and disclosure. Shame was not identified as a theme in either analysis. It emerged only on returning to the data in preparation for writing, as a sudden recognition that shame was the organising frame for everything that had already been coded. Its invisibility across two rounds of analysis – and its abrupt emergence at the point of writing – is itself consistent with shame’s characteristic tendency towards concealment, and with the particular difficulty of naming shame even for an insider researcher who shares the experience under study.

P2 had chosen to conduct her interview outdoors, in her garden, to avoid being overheard inside the house by her co-habitants. What followed was, at the time, a comical start – P2 repeatedly scanning for a rustling sound in the undergrowth, moving around the garden to find privacy, before the first author laughed together with her about the level of “general paranoia and persecution” the scene conveyed:

> “Oooh… um, it’s kind of… hold on a second… I think it’s em… hello is anyone there? No, I think it’s the next door neighbour’s dog who em… who em… oft… is anyone there? Is that Naughty Dog? No I think it’s animal it’s not a person it’s just like hearing rustling and I can’t… it’s just I keep hearing weird rustling sound in the undergrowth between our garden and the next garden I’m gonna just go slightly more over here… it’s all going in the transcript isn’t it, the general paranoia and persecution… As as as as a theme we can put that as a theme.” (P2)

This exchange was noted in the initial analysis as evidence of a very real concern for confidentiality and the risks of being identified as autistic – more pronounced in P2 than in other participants. It was not read as shame. The word does not appear in the analytical notes from that session.

The impact of this scene when applied to academic audiences was subsequently tested: with P2’s permission, two neurodivergent actors recreated it from the transcript and voice recording for conference presentations. The paralinguistic data was sufficiently rich to be performed.

On returning to the recording and reviewing it through Retzinger’s framework for identifying shame in discourse, the garden scene reads differently. The choice of location is itself a shame behaviour – concealment enacted before the interview begins. The fragmented speech (“em… who em… oft”; “as as as as”) is consistent with stammering and irregular rhythm as disorganisation of thought under shame states. The laughed words – P2 naming her own “paranoia and persecution” with laughter – are what Retzinger describes as laughed words that mask shame or embarrassment. The deflection into meta-commentary (“as a theme we can put that as a theme”) is distraction: a topic shift away from feeling. Shame was not absent from the first analysis, instead, it appeared woven throughout the interaction in ways that had not previously been recognised.

The barriers described in the sections that follow are not primarily logistical. They operate with the force they do because they are shame mediated. Autism had not merely been stigmatised externally. For these participants – who had trained and practised in the same environment that produced those stigmatising attitudes – the negative valence of autism had been internalised before it could be consciously identified or resisted.

### Sub-theme 1a: Overt shame in discourse

Shame manifested across the dataset in both overt and bypassed forms. Retzinger distinguishes between overt shame – visible, bodily, immediately identifiable – and bypassed shame, which operates below conscious awareness and surfaces through concealment, deflection, minimisation, or the paralinguistic disorganisation of speech when a topic is approached. Both were present.

Overt shame was most clearly visible in P3’s account of presenting a patient to a specialist autism team. The trigger was not hostility but the gaze itself – being observed, by professionals whose expertise centred on autism, for signs of being autistic. Shame, as Retzinger describes it, is inherently relational: it requires a perceiving other. The specialist autism team provided exactly that.

> “I then presented a patient online at one of their MDTs… I feel awful saying this because they were a very nice team but it was deeply unpleasant being watched for signs of autism.” (P3)

The aftermath was visceral:

> “It was so bad that as soon as I finished… I had to close the laptop and I couldn’t stop crying for about 20 minutes… it’s still tearing me up now and I have never felt like that before.” (P3)

What followed was a secondary shame – shame about the shame response itself, and what its visibility might cost:

> “Because I’m not gonna cry in front of people… ’cause then my credibility will be lost… not really coping… she’s not really a good doctor because now she’s crying online.” (P3)

And the resolve to present to the same team again – not withdrawal, but armoured return:

> “I’m not going to lose the one resource I’ve got for my patients… but I will have to put some armour on first.” (P3)

Three layers of shame in a single encounter: being seen as autistic, the emotional response to being seen, and the shame about that response. The armour P3 describes may function less as strength than as shame management.

P2 named humiliation explicitly when she learned that a lifelong friend had assumed she knew that she was autistic – but she had been the last to know this about herself.

> “When she said that I felt a bit like oh gosh… so she’s known this behind my back and not said anything and then I felt a bit like humiliated… maybe other people would know… [I] felt a bit thick being a psychiatrist who’s had seven years of therapy who would diagnose and recognise autism in other people… For me not to realise that until like 40 was a bit like… whoa, that’s ridiculous.” (P2)

The self-attack – *felt a bit thick* – alongside the social exposure of being known before knowing oneself, captures the relational core of shame: the self, seen through another’s eyes and found deficient.(26)

P2’s retrospective shame about her prior teaching was expressed not through a statement but through repetition – a paralinguistic pattern Retzinger identifies as fragmented, dysrhythmic speech under shame states.

> “I’m really embarrassed about the teaching I used to do about autism which was awful… awful… awful… awful.” (P2)

The shame of being seen – observed before one has chosen to be visible – was present in P5’s account of being observed stimming by a colleague after a stressful clinical situation, having believed herself unobserved. The colleague’s subsequent comment – framing what they had seen as excitement about the weekend – left the encounter unresolved, its meaning deliberately ambiguous.

> “After a very stressful clinical situation I was basically stimming really obviously… all the way back to the car… funny facial expressions and stuff, thinking that no one would be near… and then the next week one of my colleagues said ‘I saw you… you looked all excited about the weekend’…did it actually look like that or are you just saying that to see how I’d respond?” (P5)

P5 had not explained what she was actually doing. When asked directly about barriers to disclosure, she replied: *“I think fear of judgment really… of not being believed because of all the experience I’ve seen at work of so many other colleagues being so negative.”* (P5)

### Sub-theme 1b: Bypassed shame in discourse

Bypassed shame was more pervasive. Consistent with Retzinger’s observation that in bypassed states the self remains focal in awareness but is not consciously labelled as shame, P2’s description of her pre-recognition experience offers the clearest articulation:

> “I didn’t have a word for what it was, but it was definitely like this massive sense of insecurity.” (P2)

P4 described the physical register of overhearing colleagues dismiss reasonable adjustments – the moment when concealment collapsed briefly into felt experience:

> “It hurts me inside… what do you think about my reasonable adjustments when you are saying those things?” (P4)

For P6, shame carried an explicitly cultural dimension – not merely institutional but familial, intergenerational, and cross-cultural. The prospect of a disability diagnosis was not only a professional risk but a social one, extending to family honour.

> “So much of the cultural trauma that I carry from back home… revealing a disability will not be easy for me in any language in any country.” (P6)

And P3, reflecting on the cumulative cost of professional adaptation across a career, named a chronic shame state without using the word:

> “I’ve spent my career apologizing for both my strengths and my areas of challenge… which is the most depressing thing ever.” (P3)

Concealment itself – two separate lives, the managed secret, the garden interview, the stimming observed and not explained – functioned as ongoing shame management. These were unlikely to be merely strategic choices, but instead the constant, unrelenting impact of shame.

### GET 2: Origins of shame

Shame was not only present in participants’ current professional lives. Across multiple participants, childhood memories surfaced that showed shame had been seeded long before professional training compounded it: messages absorbed from family, peers, and the social environment about being fundamentally wrong.

### Sub-theme 2a: The self as defective

Before tracing the sources through which shame accumulated, it is worth noting a pattern that runs through all of them: the near-identical verbal formulation through which shame was expressed across participants. Something different, something wrong. Something wrong with me. Something wrong with her. What’s wrong with you.

> “Knowing there was this unnameable thing about me that made people not like me.” (P2)

> “I don’t think my parents would have wanted to think that there’s anything wrong with me.” (P2)

> “There does seem to be this sort of sense that you can’t be OK and be autistic.” (P3)

> “Why would you advertise that you’re in some way weird.” (P3)

> “She needs to see a psychiatrist… there’s something really wrong with her.” (P6, recounting a family response in childhood)

> “I look back on some of my gaffes with horror.” (P7)

> “I remember that feeling… when I was tiny… being different… why I wasn’t like other children.” (P5)

> “There’s something different about me.” (P4)

The phraseology does not vary substantially across participants, contexts, or decades. It appears in memories of early childhood and in accounts of recent professional encounters. It surfaces in the voices of parents, peers, colleagues, and internal self-monologue. This verbal consistency is itself a finding. That so many participants reached for almost identical language, independently and across different lives, suggests not coincidence but a shared experience: the same message was transmitted to these people, and they encoded it in the same words.

One participant did not. Following years of bullying in school, he decided at around thirteen or fourteen: “I’m just gonna be bloody different… I’m gonna do things that I want to do and that’s that” (P1). This is not the resolution shame typically produces, which is concealment. It is shame met with defiance; difference reframed as chosen rather than imposed. That this private act of self-determination did not translate into professional disclosure decades later likely speaks to the distance between personal identity and institutional safety.

### Sub-theme 2b: Family and early messages

The earliest sources of shame were familial. For several participants, the family home transmitted a clear message about the requirements of social acceptance – that difference was tolerable in private but inadmissible in public.

> “You can be as dysfunctional as you want when other people are not around but when you have outsiders looking in you need to behave normally.” (P6)

P6’s account of childhood described a life organised around unreachable social expectations whenever guests were present. Her mother experienced her daughter’s difference as a personal and social disgrace:

> “My mother… it’s a personal insult to her to say that her daughter was just so dysfunctional as a child… I remember my parents being very worried and showing me as a specimen to each other… what is wrong with her.” (P6)

P3’s childhood was marked by a similar message, absorbed from the implicit standard against which she was measured:

> “I was always in that category of could have done better… if she’d even tried a bit harder to be a bit more normal.” (P3)

For P5, the family message was delivered retrospectively – when she disclosed her autistic identity to her mother, the response confirmed what had always been communicated implicitly: “Thank goodness you noticed… that’s why you were being so weird all your childhood.” (P5) The relief in the statement did not undo its content.

P1 named the cumulative effect of a lifetime of suppression with unusual directness:

> “The dominant theme in my life has been suppressing everything. I have had to keep so much inside for so long.” (P1)

### Sub-theme 2c: School and peers

For most participants, school was where the experience of difference became explicitly social. The rejection experienced was not only painful but confusing – there was something wrong, but no language for what it was, and no way to correct it.

> “Knowing there was this unnameable thing about me that made people not like me… not having a word… I just knew there was something about me that people would dislike and I just didn’t know how to fix it, what to do differently, or how to be.” (P2)

P2 described a sixth-form incident in which the unspoken became explicit: a peer confronted her publicly with what had apparently been widely known: “Do you realise like everybody hates you? Like, do you realise like you have no friends?” (P2) The consequence was months of deliberate social exclusion. What was devastating was not only the cruelty but its familiarity; the same words would resurface, years later, in an MDT meeting (see below).

P1 responded to bullying through internalisation and performed composure:

> “I was quite badly bullied… and responded by turning it on myself, not really confiding. I remember consciously trying to appear imperturbable and on top of things even when anxious or upset… because actually expressing it never achieved anything. It just got me mocked or into further trouble.” (P1)

P7’s secondary school experience echoed this: “Secondary school was hell… I spent a lot of time wandering around on my own, avoiding bullies.” (P7)

P4, who had been diagnosed in childhood, compressed the contradiction of autistic childhood into a single phrase: “I was a bit of a horrid child. Very odd but also very able.” (P4)

The phrase very odd but also very able – held together with a *but* that does the work of mitigation – captures something essential about how shame and capability coexist in this sample. The ability does not cancel the oddness. It qualifies it. The shame is not erased by professional success; it is carried alongside it, managed, concealed, and periodically re-encountered in a colleague’s tone, an overheard comment, a rustling in the undergrowth.

### Sub-theme 2d: Generational shame

Several participants had autistic children. The recognition of autistic traits in their children was not straightforward; instead, it was accompanied by the awareness that the same environment which had produced their own shame was waiting for the next generation.

P5 described three children with different relationships to their autistic identity. One had disclosed at school and was managing: “She masks massively and really tries to be normal… she’s done really well and she’s got friends.” (P5) The description – masking as an achievement, normality as a goal, friends successfully acquired – carried within it the same logic that had shaped P5’s own concealment. A second child wanted no social contact at all. A third had done what the data suggest many autistic adults find difficult: she had named it.

> “She still says ‘I don’t want to be autistic’… so we’re doing a lot of… inspiring people, going through people historically and currently who have been good advocates, scientists, people who did good in the world who are autistic.” (P5)

’I don’t want to be autistic’ is not a simple statement of distress. It is shame made conscious and spoken aloud – consistent with Retzinger’s observation that bypassed shame characteristically resists naming even by those experiencing it. P5 was actively attempting to interrupt the transmission, to provide the counter-narrative she had not had herself. The effort was conscious, deliberate, and undertaken with the knowledge of what the alternative looked like.

P7 recognised the same traits in his own children:

> “One of them is… uncomfortably similar. Uncomfortably, because I can see it being a problem.” (P7)

The word uncomfortably appears twice. What it holds, implicitly, is the full weight of a life spent managing difference in an environment that did not accommodate it. To see it again in a child is to know exactly what that child is likely to face.

For P6, the operation of shame was visible in a different form: relief. Her son was known to be autistic. Asked if her daughter was also autistic, she described having seen nothing to concern her: “So far, I haven’t seen anything in her. She seems quite alright.” (P6) The relief is itself notable. Autism is experienced as something a child might fortunately escape. The relief registers shame’s weight by measuring what it feels like when that weight is – provisionally – absent from the next generation.

The family responses to disclosure carried their own shame signatures. When P3 told her family, the reactions were layered. Her mother’s response combined love with the instinct for concealment that shame produces:

> “We love you very much and I won’t be telling anyone… I don’t need to tell anyone… and no, thank you for the offer but I don’t really want to know any more about it.” (P3)

The love was real. The shame operating within it was also real. The “I won’t tell anyone” perfectly mirrors the professional concealment documented throughout this study, but located now within the family, where the revelation was offered in trust.

From other family members, the response to disclosure was disbelief grounded in the stereotype – expressed as a kind of compliment, but functioning as a denial:

> “We’re very surprised… that’s the last thing we’d think of for you.” (P3)

P2 encountered the same response from a close nursing colleague:

> “Oh no, but you’re really warm and you understand other people… I wouldn’t have thought that about you.” (P2)

Both responses locate the autism in the stereotype and find the person in front of them incompatible with it. The effect is to leave the disclosure unvalidated – neither believed nor disbelieved, but redirected. Shame does not require explicit rejection to be maintained; the gentle refusal to recognise is sufficient.

P3 articulated the broader stakes:

> “If only I’d known then… if only other people had known… if only we’d known.” (P3)

The triple repetition, characteristic of grief rather than regret, names what generational shame costs. Not just the individual who carries it, but the decades in which it is carried before any reckoning is possible.

P4 offered a different vantage point. Looking at late-diagnosed adults in the broader autistic community, she named what their childhoods had produced:

> “All this internalised ableism… their experience of growing up and having no clue why everything is a struggle.” (P4)

The internalised ableism P4 describes operates through shame; absorbed so thoroughly that it presents as self-knowledge rather than as something externally imposed.

### GET 3: Shame in professional life

Shame accumulated in childhood and adolescence was not left behind on entering professional life. Across several participants, the professional environment replicated the fundamental dynamic of the preceding decades: difference detected, responded to with exclusion or hostility, and met with the same imperative towards concealment.

P2’s experience in an MDT meeting years into her career returned her precisely to the vocabulary of the sixth form:

> “This social worker said to me: ‘Do you realise how much everybody hates you? Everybody hates you’… I was like, jeepers, it’s like a story of my life. And I still couldn’t figure out like why such venom.” (P2)

P1 had been told by senior mentors that he had perhaps made the wrong choice of career. The message was delivered reasonably, even kindly, but it landed where shame lands: on the question of whether one fundamentally belongs.

> “It was also apparent that I was regarded as odd by some of my mentors who did not like me… and at one point there was a conversation about whether I’d made the right choice of career.” (P1)

P3 described the experience of a career spent reading signals from an environment that could not articulate what it found unsatisfactory:

> “People kind of go a bit ‘ooh’ at you and they’re giving you signals that you’re weird… there’s something that you’ve said or you’ve done and nobody will say what… you can’t put your finger on it however hard you try, you never really get to an answer… so you just hide.” (P3)

The hiding is shame’s solution. It is also its perpetuation.

### Sub-theme 3a: Lack of knowledge, myths and misconceptions

A foundational feature of the professional environment, underlying both the shame responses described above and the barriers described below, was the pervasive lack of current knowledge about autism among psychiatric colleagues. Participants described colleagues operating from a stereotypical model of autism – male, childhood-onset, intellectually disabled – that bore little relation to the autistic identity they held.

> “That’s still their schema… the kind of pattern recognition, so when they think autism they think this little white boy with their trains literally lining them up and rocking and doing weird things with their hands… the mind blindness thing… maybe slightly impaired visual acuity when it comes to mind reading would be a better description than blindness… it’s a very powerful statement that implies that you completely can’t see something which I don’t think [is] accurate.” (P4)

> “I’ve had zero training, unless you do Learning Disability you don’t… get training in [autism]… maybe people will now… the next generation, but my generation certainly didn’t.” (P5)

This perceived ignorance was not confined to colleagues. Several participants described having held the same views before recognising themselves as autistic, and experienced significant shame on reflection.

> “I had those views five or ten years ago… I’m really embarrassed about the teaching I used to do about autism which was awful… awful… awful… awful.” (P2)

The consequences of colleagues’ perceived ignorance extended beyond missed diagnoses to the active dismissal of those who did disclose. P3 described presenting to a specialist autism team and encountering astonishment that an autistic doctor could exist.

> “The CAT [Community Autism Team] were very surprised… they were surprised that any autistic doctors existed.” (P3)

For P6, the ignorance manifested as trivialisation – the reduction of a diagnostic category to a cultural fad, heard in passing comments from colleagues.

> “Everybody is getting diagnosed… everybody has like… ‘a pinch of autism’… it’s very demoralising to hear that.” (P6)

This lack of knowledge and the stereotypes it sustained created the conditions within which shame could take root and persist across professional careers. Its operation is described throughout the sections that follow.

The stigma, disbelief, and anticipated loss of credibility described below were not expressions of deliberate hostility but emerged from an environment in which the reality of autistic professionals at senior levels remained largely invisible and poorly understood.

### Sub-theme 3b: Professional identity and credibility under threat

Participants described pervasive negative attitudes towards autism among psychiatric colleagues, encountered routinely in clinical discussions, team meetings, and supervision. Witnessing these attitudes was directly linked to concealment.

> “So many colleagues that I’ve been doing mental health act assessments with… their attitudes [towards autistic patients are] horrific… I’m really shocked by their attitudes, by how different they are to my own… I guess that’s a massive part of [not disclosing] when I hear those sort of attitudes.” (P5)

> “A colleague talking about one of our more difficult colleagues… ‘I’m sorry but she must be autistic… do you not think… I mean the way she behaves’… and the tone was not nice.” (P2)

The anticipated response to disclosure was disbelief, minimisation, or pathologisation. All participants had either experienced or feared being disbelieved.

> “Still a lot of people [think] either it’s extremely mild, so mild to the point that it doesn’t matter, or somebody clearly did not do a good job at the assessments if they have given you that label, or if you really have that then you are probably in the wrong job.” (P4)

> “It’s difficult when I hear colleagues all the time… some more experienced and respected… saying this person can’t be [autistic]… that person can’t be… it’s just a difficult culture to be believed in.” (P5)

Loss of professional credibility was a near-universal concern. For most, this centred on the perceived incompatibility of an autism diagnosis with the requirements of psychiatric practice.

> “I feel like disclosing will reduce my credibility and being able to validly diagnose others easily… so I didn’t want to.” (P2)

> “Because of the theory of mind things people would think… how could they also be a psychiatrist… how would they possibly understand what their patient’s thinking… even though I know that’s not true, people might think that… that would be a definite barrier.” (P2)

A distinct credibility threat concerned being suspected of over-investment in autism due to personal or familial connection. P3, P6, and P7 all described this.

> “In the beginning they used to [think]… ‘you see autism everywhere because your kid has it’… but that was another deterrent… because if I am autistic obviously I’ll see autism everywhere if I’m already seeing it as a mother.” (P6)

For P7, simply being seen differently was itself the threat: “Instead of seeing me as a non-autistic psychiatrist they would now see me as an autistic psychiatrist… they wouldn’t see me as the same [name].” (P7)

P6 drew a specific distinction between psychiatric colleagues and other clinical staff – it was not the broader clinical environment she experienced as unsafe, but psychiatry specifically.

> “It’s difficult to explain that I don’t feel safe… will things remain the same if I do come out with the diagnosis… the psychiatric team will not behave the same… the rest of the mental health nursing team probably would embrace it… only the psychiatric team… I don’t feel safe with them.” (P6)

### Sub-theme 3c: Diagnostic criteria and internalised ableism

The most striking illustration of internalised ableism grounded in the diagnostic criteria came from P2, describing an encounter at a mother and baby group before she had recognised herself as autistic:

> “I remember meeting someone who disclosed to me at a mother and baby group that they’re autistic… I remember thinking how can this person be a mother if they’re autistic… how are they going to get their child’s state of mind… is that actually emotionally safe for this person to be a mother… I remember thinking that and if I was thinking that as a doctor with a training in psychiatry there’s no way other psychiatrists are not gonna think that about me.” (P2)

P2 had held the ableist view herself, recognised it retrospectively as unfounded, and simultaneously understood that she was now its object. The internalisation of deficit framing was not merely an abstract barrier – it was a mirror in which participants saw the prejudice directed at autistic people, and by extension, at themselves.

The diagnostic criteria reinforced this dynamic. Their deficit-based language made self-recognition difficult and created a cognitive dissonance between the clinical picture of autism and participants’ reality as successful professionals.

> “The diagnostic criteria are just so negative… I’m the opposite of that… how could that possibly be me?” (P2)

> “You read all those papers about those horrible autistic people having no empathy and no theory of mind… is that really me? So if it’s not me, does that mean that I’m not actually autistic, or… this paper is actually very wrong? So there’s a lot of cognitive dissonance going on.” (P4)

> “The idea that actually to have a disorder like autism you must have a functional impairment… and therefore by definition there’s potentially something wrong with you doing your job.” (P1)

A further anticipated consequence – the fear of being seen through a deficit lens once disclosed – added a forward-looking threat.

> “If they had a label for it… everything I would do would be pathologized… I think people would be looking out for me to be rigid and difficult even more than they were already.” (P2)

> “Any kind of shortcoming is that gonna be attributed to that… rather than make some allowances for it, it’s gonna be like… oh this is not ever gonna get better, it cannot be fixed, is that person in the right role?” (P4)

### Sub-theme 3d: Navigating the environment – individual strategies

In response to this environment, participants described a range of strategies to manage information and control risk. Even within well-disposed teams, the perceived ignorance documented above generated responses that reinforced concealment. P3 described disclosing to a team she described as genuinely supportive – and encountering not hostility, but silence.

> “My team are very nice, very supportive, very open minded… you couldn’t wish for a nicer team but what it mostly produced was uncomfortable silence… purely because I think they didn’t know what to say… because they’ve got no frame of reference… I kind of wished that they knew a bit more.” (P3)

Loss of narrative control was a central concern – once disclosed, information could not be retrieved.

> “Right now it’s my secret… I can tell who I want and tell them to be discreet… pick and choose who I want to trust with that information and not have it as a wider thing.” (P2)

> “I don’t think they’ve got any idea how that could go once I basically lose control over my personal information… errors are made… we can’t be naive about this.” (P3)

For at least one participant, avoiding formal diagnosis was itself a navigation strategy – a way of preserving ambiguity that precluded the obligation to disclose:

> “That’s the part of me that’s like I don’t actually want this adult reassessment and a piece of paper that was gonna say it because then will I feel more obliged to tell people and… how is that gonna land?” (P4)

Witnessing the consequences of disclosure for others reinforced caution. P3 described having seen colleagues experience a shift in how they were regarded – not through malice, but through a well-meaning institutional response that nonetheless changed the relationship irreversibly.

> “Having seen what’s happened to some other people… a sort of shift in attitude towards them by other senior doctors… I don’t want to put myself in that position unless I have to… people were well-meaning, nobody was being horrible, but still it went massively awry.” (P3)

The potential for well-meaning responses to escalate was itself a deterrent – occupational health, HR, tick-box processes, all intended as support, all experienced as loss of control.

> “People will very well-meaningly want a meeting… they’ll suggest it would be a good idea if occupational health know… then they’ll suggest it’s a good idea if HR know… well-meaning, trying to do the right thing but that could massively snowball.” (P3)

A significant psychiatry-specific barrier was the professional cultural norm against self-disclosure in clinical settings, which extended naturally to the question of autistic identity.

> “While all have quirks and eccentricities, open discussion about what’s going on inside one’s head is problematic.” (P1)

> “Other than… ethically… sharing information about myself which you don’t really do in psychiatry… I think I’d feel quite confident in saying to patients yeah, I get this, I’m the same… in the way that I absolutely wouldn’t with colleagues.” (P5)

The absence of openly autistic senior role models both reflected and reinforced non-disclosure, creating a self-perpetuating cycle in which the visibility needed to make disclosure safe was itself prevented by the absence of that safety.

> “Most of the senior people who are doing well don’t disclose.” (P3)

> “It might be [that] there’s a point in the future where I feel more able to come out… more of a positive role model potentially” (P5)

### GET 4: Shame as override – why knowing the benefits isn’t enough

Participants were not unaware of the potential benefits of disclosure. Across the dataset, personal, professional, and patient benefits were articulated clearly and without ambivalence. Yet awareness of those benefits did not translate into action. That gap – between knowing and acting on that knowledge – is consistent with shame as the operative mechanism. Shame appears to not only create barriers; it renders the incentives insufficient. The capacity to act on what one knows is suspended by the same force that makes the knowing painful.

### Sub-theme 4a: The benefits of visibility – present but insufficient

Participants could identify concrete benefits of disclosure: patients who share an autistic identity with their psychiatrist make and keep appointments; teams function better when difference is named rather than managed around; visible senior autistic psychiatrists create the conditions for others to feel safer. These were not abstract or idealised benefits – participants had experienced them or witnessed them in others.

> “The only real benefit I can see is… say if I consider myself on the other side and I’m a service user I definitely would choose somebody who’s neurodivergent instead of somebody who’s neurotypical and if I disclose this is the benefit the patients, the service users would receive… that they will be able to choose.” (P6)

> “So that’s probably why I kind of end up with a lot of them on my caseload because it’s working… the kids are coming to appointments and it’s working.” (P4)

> “More of us coming out as autistic and speaking up… we’ll gradually make that feel a bit safer.” (P3)

> “The more people who disclose the more helpful it is to those who would actually improve their professional lives if they disclosed… whose colleagues might get on better with them if they had a diagnosis.” (P7)

A further benefit concerned sharing of clinical knowledge itself. Autistic psychiatrists described being unable to challenge stigmatising attitudes or correct misunderstandings about autism in the absence of disclosure – and recognised that their capacity to do so derived directly from lived experience:

> “Conversations we have every day which I find very problematic and I’m not able to flag up and it’s very difficult… the way stigmatising language was used or people just lacked the understanding and I wasn’t able to correct them… these are the things which I really sometimes struggle to make them understand… maybe I am only aware of these things because of lived experience otherwise I might have been at the same place where they are.” (P6)

The benefits were visible. They were not, however, sufficient to overcome the shame-mediated barriers documented in the preceding sections.

### Sub-theme 4b: The advocacy pull and its cost

Several participants felt a pull towards advocacy – towards using their position to make things better for other autistic psychiatrists alongside autistic patients. This pull was genuine and, in some cases, had already begun to manifest. But it carried its own shame-inflected cost.

> “Down the line a bit it would be really good to think more about if there’s anything I could do to contribute… but I don’t think now is the time so I’ll come back to you in the future… for me to get involved that would involve me potentially coming out a bit more and feeling more confident about all of that, which I don’t feel ready for yet.” (P5)

> “It’s just a simple we’re all autistic doctors being out… we can achieve great stuff… autism actually gives us strength that other people often don’t have… it’s fantastic for the population in general to see people doing so well.” (P5)

> “If you’re in a minority group you always are expecting to do the heavy lifting here… rather than the onus being on the majority to do the right thing.” (P4)

> “It’s always gonna come at a cost and I guess that’s where people need to kind of balance it and do what is right for them.” (P4)

> “If in doubt it’s easy to do nothing so it’s easier to think about it than to actually disclose… I’ve been thinking about this… it struck me might actually helpful to autistic doctors… if I said I’ve got autism.” (P1)

The advocacy pull was present alongside the recognition that acting on it required disclosure, and that disclosure remained unsafe. The result was not indifference but suspended action – wanting to contribute, knowing what it would cost, and not yet being able to pay the price.

### Sub-theme 4c: Reasonable accommodations as anticipatory shame

Participants were legally entitled to reasonable accommodations. Several had benefited from them, formally or informally. P4 had negotiated a predictable schedule, her own office, and a dedicated secretary through Access to Work [a UK government disability support scheme]. P7 had managed informally for years: “A lot of my job went well because I had a secretary who was a nanny.” (P7) P3 had made her own accommodations, working to her strengths with a line manager who had always known her.

But for P2, the accommodation itself was not the problem; it was being seen to need it – the anticipatory humiliation of being seen to need accommodations that others do not require:

> “Going through the motions of asking for accommodations would involve hoop jumping, possible ridicule and possible denial… I think I would just feel a bit humiliated asking for stuff… so I don’t think I need to actually ask for any accommodations.” (P2)

The entitlement existed. The capacity to exercise it did not. What P2 describes – dismissing a need she has acknowledged, to avoid the anticipated shame of being seen to have it – may be shame operating prospectively, foreclosing an option before it is tested. The accommodation is not refused by the institution. It is refused, pre-emptively, by shame.

P1 recognised the same dynamic without framing it as such:

> “If something ghastly happens… at that point actually looking, shining a light on how I operate, how my mind operates the way it does… might potentially be lifesaving.” (P1)

The accommodations are available but remain unasked for until a crisis makes asking unavoidable. The threshold that shame sets – catastrophe before entitlement – is its own finding.

## Discussion

This study identified shame as the overarching organising theme across autistic psychiatrists’ experiences of diagnosis and disclosure. Shame was not present as one barrier among many; it was the framework within which all barriers operated. The origins of shame in childhood and family, its transmission across generations, its operation in the professional environment, and – critically – its capacity to render known benefits of disclosure insufficient: these are unified by a single dynamic: the internalisation of a cultural message about autism as deficit, absorbed through training, applied in clinical practice, and ultimately turned on the self.

### Shame and stigma: a necessary distinction

Shame is not the same as stigma. Stigma describes external social labelling – the attribution of discrediting characteristics to a group.(31) Shame is the affective component of stigma: what happens when that labelling is internalised, when the self rather than the environment becomes the site of the problem.(32) This distinction matters clinically and theoretically. The barriers to disclosure and diagnosis documented here are not primarily explained by stigma, that is, by what colleagues might think or do. They are explained by shame – by what participants had already come to think and feel about themselves as autistic people. P2’s inability to recognise herself in the diagnostic criteria she had been teaching for years; P3’s twenty minutes of crying after being watched for signs of autism by people whose expertise she respected; P1’s lifelong suppression as the dominant life theme: none of these are straightforwardly external. They are the residue of a stigmatising environment that had been thoroughly absorbed.

The conflation of shame and stigma is illustrated in a recent qualitative study of autistic parents of autistic children, in which barriers to research participation were attributed to ‘self-stigma’ and ‘internalised stigma’ – yet one participant described the prospect of being videoed as potentially ‘a shaming experience,’ and another feared discovering ‘I’ve been doing something detrimental to my kids without realising.’(33, 34) These are not responses to external labelling; they are anticipatory shame – the dread of being seen and found deficient. That the author, herself autistic and deeply aware of stigma, did not initially recognise these as shame responses is consistent with shame’s characteristic resistance to naming.

Alternative interpretations were considered throughout the analysis. Fear of discrimination, anticipated stigma, minority stress, and rational career protection are all present within the dataset and explain significant aspects of the findings. However, none adequately accounts for the full picture. Fear and stigma explain avoidance of external threat, but not the inability to recognise oneself in diagnostic criteria one has spent years applying to others; not the emotional intensity of being observed by an autism assessment team; not the avoidance of accommodations before any institutional refusal has occurred; not the recurrence of structurally similar patterns across childhood, family life, and professional domains separated by decades. Shame provided the most parsimonious account of these otherwise disparate observations.

This finding is consistent with Dolezal and Bynum’s argument that shame is pervasive in healthcare and remains unnamed and unaddressed, contributing to burnout, depression, and impaired functioning among clinicians.(35) It extends that argument by identifying shame as operating specifically along the axis of neurodevelopmental identity within psychiatry – the very specialty most responsible for producing the deficit-based framing that generates it.

The analytical sequence – initial analysis, recognition of shame, return to data, shame found throughout – requires scrutiny. The risk of circularity is real: once the analyst went looking for shame, shame could be presumed inevitable. Three features of the analysis argue against this reading. First, and most significantly, shame was not the initial hypothesis. It was absent across two prior analytical rounds, emerging only at the point of writing – after the initial analysis was complete. If the analyst had been primed to find shame, it would have appeared earlier. Its late emergence, in the context of data the analyst knew intimately, demonstrates the opposite of confirmation bias. Second, the Retzinger framework identifies specific paralinguistic and verbal indicators that are theoretically distinct from the markers of fear, anxiety, or strategic concealment – the most plausible alternative interpretations of the data. The bypassed shame indicators in particular (stammering, irregular rhythm, laughed words, distraction, verbal hiding) are not predicted by minority stress or strategic identity management frameworks. Third, independent verification by a co-author working from transcripts, without access to the first author’s interpretive notes, provided external anchoring for the shame identification. The circularity risk was real; these features of the analysis mitigate it without eliminating it entirely.

### The self-perpetuating cycle

However, the findings indicate a closed loop that existing literature has not yet fully described in this population. Shame drives concealment. Concealment means autistic psychiatrists remain invisible to colleagues, to trainees, and to patients.

Invisibility sustains the assumption – false, but powerful – that autism is incompatible with psychiatric practice. That assumption generates further shame. As the data document, even the most supportive response to disclosure – P3’s mother’s “we love you and I won’t be telling anyone” – reproduces the logic of concealment that shame requires.

This cycle has consequences that extend beyond individual wellbeing. Disclosure enables colleagues and trainees to calibrate their clinical pattern recognition more accurately, and facilitates direct teaching informed by lived autistic experience.(23) The patient-level consequences of this underutilised resource are reported in depth elsewhere.(36)

Autistic psychiatrists who conceal their identity cannot function as role models – and the absence of role models was itself identified as a barrier to disclosure. The resulting invisibility sends an implicit message to autistic trainees, autistic colleagues, and autistic patients: that autism and psychiatric competence are incompatible. The very psychiatrists best placed to challenge that message, by virtue of their insider knowledge, are silenced by the same shame the message produces.(23)

The scale and impact of this invisibility is documented in the wider autistic doctors’ literature. In a cross-sectional survey of autistic doctors, 80% reported having worked with a colleague they *suspected* was autistic, yet only 22% had worked with a colleague they *knew* was autistic, itself evidence of the taboo against naming what is already suspected.(4) The gap between suspicion and knowledge is not a data artefact. It is shame made structural: a professional culture in which autistic identity is widely inferred but almost never confirmed. The taboo against asking is simultaneously a measure of the stigma and shame that make disclosure unsafe, and a mechanism for perpetuating both. The consequences of that gap are not trivial: having never worked with another doctor they suspected to be autistic was significantly associated with having considered suicide.(4) Role models in medical education and practice matter,(37) not as an abstract principle but as a quantifiable protection against the most serious outcomes of professional isolation.

### The override function of shame

Crucially, shame becomes dissociated from its original social source. Stigma can be intellectually processed and rejected; shame persists as an affective residue independent of that processing. This is why autistic psychiatrists who can articulate epistemic injustice, who teach about stigma, who actively embrace a neurodiversity-affirmative understanding of their own identity, still cannot disclose. The cognitive work has been done. The shame remains.

A distinct dimension of shame’s operation emerged in GET 4, and provides perhaps the strongest evidence that shame, rather than fear or stigma, is the operative mechanism. Participants knew the potential benefits of disclosure – they articulated them clearly: patients choosing autistic clinicians, teams functioning better when difference is named, trainees finding role models, the collective visibility that makes disclosure progressively safer. This was not a knowledge deficit. The capacity to act on that knowledge was, nonetheless, suspended.

Fear of discrimination, anticipated stigma, and rational career protection predict avoidance of disclosure where outcomes are uncertain. They do not predict the inability to act where participants can clearly articulate the benefits – including benefits to themselves, their colleagues, and their patients. The gap between knowing and doing is most parsimoniously explained by shame: an affect that operates at the level of internal prohibition rather than external sanction. This has direct implications for how disclosure barriers in this population are understood and addressed. Interventions framed around raising awareness of disclosure benefits, or around reducing external stigma through education, misread the underlying mechanism. Where shame is the organising force, it renders incentives insufficient before they reach the threshold of action. The most striking illustration is the finding on reasonable accommodations: participants were legally entitled to accommodations, in some cases had negotiated them formally, and yet others could not bring themselves to ask. The entitlement existed but the capacity to exercise it did not. Shame inhibited the option prospectively – not because the institution refused, but because being seen to need the accommodation was, in itself, the threat. Where stigma operates at the level of external sanction, shame operates at the level of internal prohibition. Removing the former leaves the latter intact.

### Psychiatry-specific dimensions

Several features of shame in this sample were specific to psychiatry as a professional context. The theory of mind and empathy concerns – that autistic psychiatrists could not understand their patients’ mental states – represent a direct application of deficit-based diagnostic framing to professional competence.

Participants had absorbed these concerns themselves: P2’s prior view that an autistic mother could not adequately understand her child’s emotional state was, she recognised, the same view that would be applied to her. The training that produced diagnostic skill had simultaneously produced the internalised shame that prevented disclosure.

The culture of non-disclosure that is normative in psychiatric practice – the prohibition on self-disclosure to patients or colleagues – provided institutional cover for the concealment that shame demanded. As P1 observed, open discussion about what is going on inside one’s head is problematic in psychiatry. The professional norm and the shame response were structurally aligned: psychiatry already required concealment, and shame made concealment feel necessary. These two forces reinforced each other.

### Diagnostic criteria as a structural source of shame

A specific structural contributor to shame in this sample was the diagnostic criteria themselves. The deficit-based framing embedded in DSM and ICD autism criteria – emphasising impairment, disorder, and functional limitation – creates a double bind for high-achieving autistic professionals. First, it renders self-recognition difficult: participants described a profound cognitive dissonance between the clinical picture of autism they had been taught and their own functioning as competent, credentialled psychiatrists. Second, it makes the diagnostic label itself shameful within a professional context, because receiving it formally implies the functional impairment that the criteria require – and functional impairment in a psychiatrist implies professional incompetence. In this sample, the deficit-based framing of diagnostic criteria functioned as a structural source of shame. It is not incidental that the phrase “if you really have that then you are probably in the wrong job” was widely anticipated by participants: this is the logical conclusion of applying deficit-based diagnostic criteria to a professional population. For these participants, the criteria did not merely fail to fit – they actively generated shame in those they were supposed to describe.

This has a direct consequence for disclosure. Formal diagnosis under a deficit framework does not just label someone as autistic – it labels them as impaired. In a clinical environment where impairment is understood as incompatibility with psychiatric practice, formal diagnosis may become a professional liability for these participants. The avoidance of diagnosis documented here is not irrational; it is a protective response to a diagnostic system whose framing makes the label dangerous.

### Comparison with existing literature

A recent qualitative study using template analysis identified shame as an organising force in autistic burnout among autistic adults, finding that shame shaped interpretations of functional limits, masking efforts, and help-seeking.(38) The present findings extend this in several important directions. The population – autistic psychiatrists specifically – introduces dimensions absent from general autistic adult samples: the professional identity stakes of disclosure, the presence of colleagues as potential assessors, the normative prohibition on self-disclosure, and the downstream consequences for patients. The methodology – IPA using Retzinger’s framework as an analytical tool within the interpretive process – identifies shame not only in what participants said but in how they said it: the paralinguistic indicators present across two rounds of analysis that only became visible when specifically sought.

A qualitative phenomenological study of autistic foundation year doctors identified shame as a subtheme of professional identity, emerging as one consequence of the pressures to conform and conceal.(39) In that study, shame was an endpoint – one among several outcomes of navigating a system not designed for autistic people.

Another qualitative study of autistic medical students’ experiences of peer support similarly identified shame as a downstream consequence of stigma, weaponised professionalism, and internalised ableism – described variously as a culture of silence and, simply, “a dirty secret”.(40) The present findings reposition shame as the mechanism: not where the journey ends, but the force organising the journey from the beginning. Both studies were conducted by researchers with overlapping authorship with the present paper but with independent analysis teams. In the case of Keenan et al., the lead analyst was aware of the emerging shame findings in this dataset during the analytical phase, having heard the first author present preliminary findings; the shame subtheme in that study emerged independently.

The independent emergence of shame across three separate datasets – autistic adults in burnout, autistic foundation year doctors, and autistic medical students – using different methodologies and independent analysis teams, provides external support for the present findings that goes beyond comparison. Shame is not appearing only in this dataset; it is being found repeatedly, by different researchers, in adjacent populations. The conflation of shame and stigma in adjacent literature suggests that reviewing existing work on stigma through a shame lens may be a worthwhile direction for future research.

### Implications for practice

The findings suggest that interventions framed around increasing disclosure among autistic doctors misread the problem. Disclosure may carry genuine risks; to career progression, to professional credibility, to the control of one’s own narrative, that the data document clearly and that participants navigated with sophistication. Non-disclosure in the current environment is not necessarily a failure of courage or identity acceptance; it is a rational response to real institutional threat.

The goal is not more disclosure but conditions in which the decision about disclosure – in whatever direction – can be made without shame as the primary driver. Shame-sensitive approaches, as described by Dolezal and Bynum,(35) would reduce the cost of visibility for those who choose it: environments where autistic identity is not framed as deficit, where disclosure does not automatically trigger institutional machinery, and where a psychiatrist’s competence is not retroactively reinterpreted through a deficit lens. Equally, they would involve reducing the shame that attaches to concealment, while recognising that keeping one’s autistic identity private is a legitimate and reasonable choice in conditions that currently make openness genuinely dangerous.(23)

### Shame, psychiatry, and the case for a paradigm shift

The findings point toward a specific structural solution: a shift from deficit-based to neurodiversity-affirmative approaches in psychiatric training and practice. A neurodiversity-affirmative framing understands autism as a form of human variation rather than disorder, recognises that impairment is produced at the intersection of neurological difference and an environment that does not accommodate it, and does not assume that professional competence is incompatible with an autistic profile.(41) This framing does not minimise the real and significant support needs of some autistic people; it contests the assumption that disability and professional competence are mutually exclusive. Under such a framework, the cognitive dissonance that makes self-recognition difficult dissolves: a psychiatrist who is excellent at their work and autistic is not a contradiction, but an expected and valued reality.(42)

The implications of this shift extend in two directions. For autistic psychiatrists, a neurodiversity-affirmative environment reduces the professional cost of an autistic identity, both diagnosed and disclosed. If autism does not imply impairment, disclosure does not imply professional inadequacy. The decision about whether to disclose becomes genuinely free rather than structurally coerced by shame. For the broader psychiatric workforce and patient population, autistic psychiatrists who are visible and supported, not as exceptions who overcame their “deficits”, but as practitioners whose neurocognitive profile is a legitimate variant, provide the role models whose absence was itself identified as a barrier. A neurodiversity-affirmative psychiatry does not merely benefit autistic psychiatrists.(43) It changes the epistemic environment in which all psychiatrists learn to see and understand autism, with direct consequences for the accuracy and quality of autism assessment and the care of autistic patients.(44)

The existence of autistic psychiatrists challenges the deficit-based framework from the inside. But shame-mediated non-disclosure means that challenge is largely unheard: the people best placed to contest the narrative that autism is incompatible with professional attainment are prevented, by the same shame that narrative generates, from making their case. The result is not merely a gap in the evidence. It is the active perpetuation of a model of autism that is inaccurate – sustained, in part, by the silence of those whose lives refute it. As P4 observed, *“If you were using all those criteria and have it deeply engrained in your head… what the picture of autism is… the cognitive dissonance just blows people minds”*.

### Limitations

The sample size (n=7) is small but appropriate for IPA, which prioritises interpretive depth over breadth. Participants were predominantly UK-based and recruited through a psychiatry-specific autistic professional network. Recruitment through ADI may skew the sample in two opposing directions simultaneously: towards those who have achieved sufficient comfort with their autistic identity to seek community, and towards those whose professional isolation or distress made peer connection particularly important. Those most severely affected by shame-driven concealment, and those who have not yet recognised themselves as autistic, are structurally absent – and that absence is itself theoretically significant given the phenomena under investigation.

The majority of participants self-identified as autistic without a formal diagnosis. Self-identification is a recognised practice within autistic communities, and all participants were psychiatrists with clinical expertise in autism diagnosis. The data themselves illuminate why formal diagnosis may be actively discouraged in this professional context. One participant described seeking advice from a supervisor about pursuing a formal diagnosis and being told:

> “why go for a diagnosis… you’re a psychiatrist, you’ve diagnosed yourself, what does anyone else need to tell you [that] you don’t know yourself… get a diagnosis? for who? for what?”(P2)

Acceptance within the ADI Psych group, which consists of autistic and autism-diagnosing psychiatrists, provides community confirmation of autistic identity. Nothing in the data gives us reason to question the autistic identity of any participant.

The insider methodological approach enabled interpretive access unlikely to be available to non-autistic researchers, while simultaneously introducing the risk of over-identification with participants and the possibility that aspects of shared experience may shape what is foregrounded or occluded. This tension is engaged with throughout the positionality and reflexivity sections rather than resolved. One participant was excluded following institutional review due to concerns about identification, reducing the sample from n=8 to n=7. This exclusion did not result in the loss of any group experiential theme.

The perspectives represented here are those of senior autistic psychiatrists; trainee doctors were not included. This represents a narrow subset of autistic people and does not represent the full range of autistic experience, excluding in particular autistic people with intellectual disability, higher support needs, or limited verbal communication.

### Conclusion

Autistic psychiatrists navigate diagnosis and disclosure within an environment that makes both genuinely risky. The barriers documented here are not primarily logistical but shame-mediated: sustained by the internalisation of deficit framing, reinforced by professional norms of concealment, and transmitted across generations. The response to that environment – concealment, strategic disclosure, avoidance of formal diagnosis – is rational. The problem is the environment and the culture within psychiatry. Naming shame, in research, in clinical practice, and in psychiatric training, is a prerequisite for creating conditions in which autistic psychiatrists can make decisions about their own identities freely, rather than under the organising force of shame.

## Data Availability Statement

The data that support the findings of this study are not available to share. Ethical approval (London South Bank University Research Ethics Committee; ETH2122-0128) does not support data sharing, and participants did not consent to such sharing. The data are highly sensitive: participants are members of a small, professionally identifiable group, and the interview transcripts contain detailed personal accounts of diagnosis, disclosure, and mental health that could not be adequately de-identified. Anonymised quotes are included throughout the results section to evidence the analysis and support interpretation. Researchers wishing to make a data access request may contact the corresponding author; ethical queries can be addressed to.

## References

1. Woods S. Why we need more autistic health care professionals and how to support them. Autism in Adulthood. 2026;8(1):5–13.

2. Wood R, Happe F. What are the views and experiences of autistic teachers? Findings from an online survey in the UK. Disability & Society. 2021:1–26.

3. Hawker D, Muggleton J, Henshaw E, Horne K, Hutchinson J, Little L, et al. Neurodiversity is not just for those we work with: A group of autistic psychologists write. The Psychologist. 2022.

4. Shaw SCK, Fossi A, Carravallah LA, Rabenstein K, Ross W, Doherty M. The experiences of autistic doctors: a cross-sectional study. Front Psychiatry. 2023;14:1160994.

5. Curnow E, Maciver D, Johnston L, Murray M, Johnstone-Cooke V, Utley I, et al. Learning from the Experiences of Autistic Professionals Working in Health and Education. Autism in Adulthood. 2025.

6. Lord C, Charman T, Havdahl A, Carbone P, Anagnostou E, Boyd B, et al. The Lancet Commission on the future of care and clinical research in autism. Lancet. 2022;399(10321):271–334.

7. Mottron L. A radical change in our autism research strategy is needed: Back to prototypes. Autism Res. 2021;14(10):2213–20.

8. Lewton T. Why autism pioneer Uta Frith wants to dismantle the spectrum. New Scientist. 2026 May 13. https://www.newscientist.com/article/2525037-why-autism-pioneer-uta-frith-wants-to-dismantle-the-spectrum/

9. Clarke EB, McCauley JB, Lutz A, Gotelli M, Sheinkopf SJ, Lord C. Understanding profound autism: implications for stigma and supports. Front Psychiatry. 2024;15:1287096.

10. Zhang X, Grove J, Gu Y, Buus CK, Nielsen LK, Neufeld SAS, et al. Polygenic and developmental profiles of autism differ by age at diagnosis. Nature. 2025;646(8087):1146–55.

11. Mottron L, McGuire OL, Gagnon D. Autism-ness Does Not Exist, but Autism Does. Part 1: A Critic of the “Spectrum” Position Used to Describe, Diagnose, and Research Autism, and Its Alternative. Autism Dev Lang Impair. 2025;10:23969415251404764.

12. Vincent J, Ralston K. Uncovering employment outcomes for autistic university graduates in the United Kingdom: An analysis of population data. Autism. 2024;28(3):732–43.

13. Doherty M, Johnson M, Buckley C. Supporting autistic doctors in primary care: challenging the myths and misconceptions. Br J Gen Pract. 2021;71(708):294–5.

14. Romualdez AM, Walker Z, Remington A. Autistic adults’ experiences of diagnostic disclosure in the workplace: Decision-making and factors associated with outcomes. Autism Dev Lang Impair. 2021;6:23969415211022955.

15. Romualdez AM, Heasman B, Walker Z, Davies J, Remington A. ‘People might understand me better’: Diagnostic disclosure experiences of autistic individuals in the workplace. Autism in Adulthood. 2021;3(2):157–67.

16. Evans JA, Krumrei-Mancuso EJ, Rouse SV. What you are hiding could be hurting you: Autistic masking in relation to mental health, interpersonal trauma, authenticity, and self-esteem. Autism in Adulthood. 2024;6(2):229–40.

17. Pryke-Hobbes A, Davies J, Heasman B, Livesey A, Walker A, Pellicano E, et al. The workplace masking experiences of autistic, non-autistic neurodivergent and neurotypical adults in the UK. PLOS One. 2023;18(9):e0290001.

18. Russell AS, McFayden TC, McAllister M, Liles K, Bittner S, Strang JF, et al. Who, when, where, and why: A systematic review of “late diagnosis” in autism. Autism Res. 2025;18(1):22–36.

19. Doherty M, Shaw S, Chaplin E, Baron-Cohen S. Embracing neurodiversity in medicine: insights from the history of psychiatry. The Lancet. 2026.

20. Zeidan J, Fombonne E, Scorah J, Ibrahim A, Durkin M, Saxena S, et al. Global prevalence of autism: A systematic review update. Autism Research. 2022;15(5):778–90.

21. O’Nions E, Petersen I, Buckman JEJ, Charlton R, Cooper C, Corbett A, et al. Autism in England: assessing underdiagnosis in a population-based cohort study of prospectively collected primary care data. Lancet Reg Health Eur. 2023;29:100626.

22. Abeyratne L, Courtenay K, Al-Najjar Z, Shaw SC, Odiaka V, Doherty M, et al. Autistic Traits in Doctors: A Cross-sectional Study of Doctors Seeking Mental Health Support. Research Square [Preprint]. 2025. 10.21203/rs.3.rs-7437542/v1

23. Doherty M, Henderson J, Rebowska A, Grosjean B, Kinnear M, Dakin C, et al. Two more elephants in the room: autistic psychiatrists and autistic shame. Br J Psychiatry (in press). 2026.

24. Turnock A, Langley K, Jones CR. Understanding stigma in autism: A narrative review and theoretical model. Autism in Adulthood. 2022;4(1):76–91.

25. Doherty M, Chown N, Martin N, Shaw SCK. Autistic psychiatrists’ experiences of recognising themselves and others as autistic: a qualitative study. BJPsych Open. 2024;10(6):e183.

26. Shaw SCK, Anderson JL. Phenomenological research in medical education: an overview of its philosophy, approaches and conduct. In SAGE Research Methods Cases Part Two. 2018.

27. Retzinger SM. Identifying shame and anger in discourse. American Behavioral Scientist 1995;38(8):1104–13.

28. Smith JA, Nizza IE. Essentials of interpretative phenomenological analysis: American Psychological Association; 2022.

29. Motulsky SL. Is member checking the gold standard of quality in qualitative research? Qualitative Psychology. 2021;8(3):389–406.

30. Doherty M. Autistic Shame: Reflections on an Under-Recognised Determinant of Autistic Mental Health 2026. 10.31234/osf.io/kn95m_v1

31. Goffman E. Stigma; notes on the management of spoiled identity. Englewood Cliffs, N.J.,: Prentice-Hall; 1963. 147 p.

32. Hutchinson P, Dhairyawan R. Shame, stigma, HIV: philosophical reflections. Medical humanities. 2017;43(4):225–30.

33. Heyworth M. Commentary: Addressing stigma through relational research design. Neurodiversity. 2024;2:27546330241230428.

34. Heyworth M, Tan DW, Pellicano E. “I Think I’m Really Well-Equipped to be a Parent”: Autistic Parents’ Experiences of Parenting Autistically. Autism in Adulthood. 2026:25739581261426626.

35. Dolezal L, Bynum W. Shame competence: addressing the effects of shame in health care. Lancet. 2024;404(10462):1514–5.

36. Doherty M, Chown N, Martin N, Grosjean B, Shaw SC. Autistic Psychiatrists’ Perspectives on Mental Healthcare for Autistic People: A Qualitative Study. [Preprint] medRxiv 2026. 10.64898/2026.06.01.26354595

37. Shaw SCK, Grosjean B, McCowan S, Kinnear M, Doherty M. Autistic role modelling in medical education. Education for Primary Care. 2022;33(2):128–9.

38. Clarey MM, Ireland MJ, Abel S, Brownlow C. Beyond Exhaustion: Shame, Identity Disruption, and Functional Collapse in Autistic Burnout. Autism. 2026:13623613261444797.

39. Keenan J, Doherty M, Shaw SCK. The Experiences of Autistic Doctors Transitioning into Clinical Practice in the UK: A Phenomenological Study. Teaching and Learning in Medicine. 2026:1–15.

40. Cooper S, Doherty M, Shaw SCK. The experiences of autistic medical students in relation to seeking and receiving online support: A phenomenological study. PLOS One. 2026;21(3):e0345156.

41. Pellicano E, Fatima U, Hall G, Heyworth M, Lawson W, Lilley R, et al. A capabilities approach to understanding and supporting autistic adulthood. Nature Reviews Psychology. 2022:1–16.

42. McCowan S, Shaw SC, Doherty M, Grosjean B, Blank P, Kinnear M. A full CIRCLE: inclusion of autistic doctors in the Royal College Of Psychiatrists’ values and Equality Action Plan. The British Journal of Psychiatry. 2022:1–3.

43. Shaw SC, Doherty M, McCowan S, Eccles JA. Towards a neurodiversity-affirmative approach for an over-represented and under-recognised population: autistic adults in outpatient psychiatry. Journal of Autism and Developmental Disorders. 2022;52(9):4200–1.

44. Doherty M, Shaw SC, Dakin C, Milton D, Morton H. The ADOS-2 rapport item: an unexamined assumption. The Lancet Psychiatry. 2026.

